# Ferritin-guided iron supplementation in whole blood donors (FORTE): results of a double-blind randomized controlled trial

**DOI:** 10.1101/2025.01.03.25319940

**Authors:** Jan H M Karregat, Amber Meulenbeld, Franke A Quee, Vĕra M J Novotny, Masja de Haas, Dorine Swinkels, Bert-Jan H van den Born, Hans L Zaaijer, Jos W R Twisk, Katja van den Hurk

**Affiliations:** Donor Health, Sanquin Research, Amsterdam, The Netherlands; Department of Public and Occupational Health, Amsterdam UMC, Amsterdam, The Netherlands; Amsterdam Public Health (APH) Research Institute, Amsterdam UMC, Amsterdam, The Netherlands; Department of Medical BioSciences, Radboudumc University Medical Center, Nijmegen, The Netherlands; Department of Medical Affairs Sanquin, Amsterdam, The Netherlands; Department of Laboratory Medicine, Radboud, Nijmegen, The Netherlands; University of Amsterdam, Department of Internal Medicine, Section Vascular Medicine, Amsterdam Cardiovascular Sciences, Amsterdam UMC, Amsterdam, The Netherlands; Blood-borne Infections, Sanquin Research, Amsterdam, The Netherlands; Department of Epidemiology and Biostatistics, Amsterdam UMC, Amsterdam, The Netherlands

## Abstract

**BACKGROUND:** Regular blood donation is associated with an increased risk of iron deficiency (ID; ferritin <15 µg/L). Oral iron supplementation is known to shorten iron store recovery time and could serve as a more effective alternative to extended donation intervals. We aimed to determine the optimal ferritin-guided iron supplementation protocol in terms of donor health, side-effects, donor return, and treatment adherence.

**METHODS:** In this prospective randomized placebo-controlled trial donors with ferritin levels ≤30 µg/L were enrolled in the study and randomly assigned to one of six groups, stratified for sex and age. Depending on the study arm, donors were asked to adhere to a ferrous bisglycinate supplementation protocol for 56 days, taking capsules containing 0 mg (i.e. placebo), 30 mg or 60 mg of elemental iron, either on alternate days or daily. The primary outcome was iron deficiency (ID) at 56 days follow-up, while secondary outcomes included low ferritin (≤30 µg/L), low Hb (≤135 g/L for men and ≤125 g/L for women), iron deficiency-related symptoms, side effects, treatment adherence, and evaluations of all outcomes at 122 and 180 days follow-up. This trial is registered in the Dutch trial registry (NL73283.018.20).

**FINDINGS:** Of the 2,052 donors who provided their informed consent, 830 donors (464 women) had ferritin levels ≤30 µg/L and were included in the trial. Compared to placebo, all iron supplementation groups exhibited similar significantly lower odds of ID at 56 days, with odds ratios (OR) ranging from 0.60 (95% CI [0.55–0.66]) to 0.65 (95% CI [0.59–0.72]). Similarly, for low Hb, the OR’s ranged from 0.74 (95% CI [0.64–0.86]) to 0.80 (95% CI [0.68–0.93]) For low ferritin, 60 mg of daily iron supplementation yielded an OR of 0.52 (95% CI [0.47–0.57]), markedly lower than those of other protocols, which ranged from 0.61 (95% CI [0.55–0.68]) to 0.82 (95% CI [0.74–0.91]). Iron supplementation did not result in any significant differences compared to placebo in ID-related symptoms, gastrointestinal side effects, intention to return to donate, or treatment adherence.

**INTERPRETATION:** In regular donors, 60 mg iron supplements taken daily was shown to be the most effective strategy for mitigating ID, low Hb, and especially low ferritin without introducing gastrointestinal discomfort or changes in the donors intention to return to donate. Ferritin-guided iron supplementation is an effective iron management strategy as an alternative or supplement to extended donation intervals.

## INTRODUCTION

Worldwide, over 100 million whole blood donations are made each year, providing a critical resource for life-saving transfusions and treatments.^1^ This invaluable supply depends on the generosity of millions of blood donors. Regular whole blood donation is associated with loss of iron and an elevated risk of developing iron deficiency (ID; ferritin <15 µg/L) and low haemoglobin (Hb) levels, potentially threatening donor health and donor availability.^2^ Blood services primarily focus on monitoring Hb levels to prevent anaemia and ensure the quality of blood products. European legislation mandates that the donor eligibility threshold for Hb levels among male and female whole blood donors be set at >135 g/L and >125 g/L, respectively. However, Hb levels do not accurately reflect the body’s iron stores, and anaemia can be a late consequence of ID. Consequently, ferritin-based deferral policies are increasingly common among blood services as a means to monitor donors’ iron stores and prevent ID.^3,4^ We have recently shown that ferritin-guided donation intervals significantly improved Hb and ferritin levels in whole blood donors. However, donor availability was negatively impacted due to longer donation intervals and decreased donor return.^5^

An alternative approach to enhance iron and Hb recovery in regular whole blood donors is post-donation iron supplementation, which is already used by blood services in the USA, Denmark, and Finland.^6^ In a randomized clinical trial, Kiss *et al.* previously showed that iron supplementation for 56 days following blood donation can lead to fully recovery of ferritin to pre-donation levels^7^. However, the optimal supplementation strategy for whole blood donors to improve iron stores remains unclear. Studies examining supplementation of iron in whole blood donors examined dosages ranging from 19 mg/day to 240 mg/day.^8,9^ However, studies have shown that lower dosed iron supplements (19 mg) may be equally effective compared to higher dosed supplements (38 mg) in improving ferritin levels.^9^ Additionally, recent studies suggest that alternate days, rather than daily iron supplementation, may improve iron uptake in the intestine.^8,10,11^ This may potentially reduce gastrointestinal discomfort associated with iron supplementation, but also impact treatment adherence.

At present, the optimal dosing strategy and intake frequency of iron supplementation for improving iron recovery, minimizing gastrointestinal discomfort, and ensuring treatment adherence remain elusive, and its efficacy in both male and female whole blood donors has yet to be established. Therefore, we aimed to evaluate the effects of varying iron supplement dosages and intake frequencies on ID and ID-related symptoms in whole blood donors, compared to placebo supplementation.

## METHODS

### Setting and design

We used a randomized double-blind placebo controlled trial design to study the effect of different iron supplementation dosages and dosing frequencies on ID and related symptoms using the Sanquin blood services infrastructure of the Netherlands. Sanquin is a not-for-profit organisation and the only organisation in the Netherlands with a legal permission and obligation to collect, process, and provide blood products for transfusion and the production of blood-derived medicinal products. Eligibility for donation is evaluated through a donor health questionnaire and onsite assessment prior to every donation. Donors can donate when they are in good health, at least 18 years old, and not at risk for any transfusion-transmissible infectious diseases. Hb levels should be >135 g/L and >125 g/L for men and women, respectively, and are measured in capillary blood (HemoCue 201, Angelholm, Sweden). In the Netherlands, donors with Hb levels below the cut-off are deferred from donation for at least 3 months. Additionally, starting November 2017, Sanquin has implemented a ferritin-guided donation interval policy. Ferritin levels are measured in serum (Architect Ci8200, Abbott Laboratories, Illinois) post-donation in newly registered donors and at every fifth whole blood donation. Donors are deferred for 6 or 12 months as a consequence of ferritin levels between ≥15-≤30 µg/L or <15 µg/L, respectively.

The protocol of the FORTE trial has been previously described in detail.^12^ In brief, between August 2021 and January 2023 donors were invited via email and provided with the baseline questionnaire if their ferritin levels were measured at their next donation. Donors were excluded from participation when they had taken iron supplements regularly in the 3 months prior to enrolment. If their ferritin level was ≤30 µg/L, and thus deferred for at least 6 months, donors were randomized into one of six groups based on iron supplementation dosage and intake frequency: placebo daily (PD), placebo alternate day (PA), 30 mg daily (30D), 30 mg alternate day (30A), 60 mg daily (60D), and 60 mg alternate day (60A). Donor were asked to adhere to the intake protocol for 56 days, and were subsequently invited for their first follow-up visit. During the post-supplementation period, donors were invited for follow-up visits 2 and 3 at 122 and 180 days, respectively. This study is performed according to the Declaration of Helsinki and Good Clinical Practice guidelines. The Medical Research Ethics Committee Amsterdam UMC has approved the study protocol (trial ID NL8590) and is registered in the Dutch trial registry (NL73283.018.20).

### Intervention and timeline

Participants received blister packs containing either thirty capsules for alternate day intake or sixty capsules for daily intake, along with instructions regarding their prescribed intake frequency. The capsules contained placebo (0 mg elemental iron), low-dose iron (30 mg elemental iron), or high-dose iron (60 mg elemental iron). The participants were instructed to strictly adhere to the prescribed intake regimen for a duration of 56 days, and to refrain from eating dietary product which are associated with reduced iron uptake (e.g., dairy product, coffee, and tea) shortly before or after supplementation. After 56, 122 and 180 days, participants were scheduled for follow-up visits. Participants were instructed to abstain from consuming any other iron-containing dietary supplements throughout the entire duration of the study.

### Outcomes

The primary outcome of this study was the presence of ID, defined as a serum ferritin <15 ug/L at 56 days follow-up. Secondary outcomes included presence of low ferritin, defined as serum ferritin 15-30 µg/L, and low Hb, defined as Hb levels ≤135 g/L for men and ≤125 g/L for women. Other secondary outcomes included gastrointestinal side-effects evaluated by the Gastrointestinal Symptom Rating Scale^13^; ID-related symptoms including general health (*36-Item Short-Form Health Survey*^14^), fatigue (*Fatigue Assessment Scale*^15^), cognitive failure (*Cognitive Failure Questionnaire*^16^), restless legs syndrome (RLS) (*Cambridge Hopkins Restless Legs Syndrome Questionnaire*^17^); intention to return to donate (*Theory of Planned Behavior*^18^); and treatment adherence. Treatment adherence was evaluated by counting the number of returned capsules at 56 days and, if not available, through the data from the treatment adherence application MedApp (MedApp Nederland B.V., Eindhoven, The Netherlands, https://medapp.nl), which has been described previously^12^.

### Statistical analysis

Baseline descriptive characteristics are described for all trial participants, stratified by trial arm. Continuous data are expressed as mean ± standard deviation, or as median with interquartile range when not normally distributed. Categorical data are presented as proportions.

We conducted Linear Mixed Models (LMM) analysis for continuous outcomes and Generalized Estimating Equation (GEE) analysis for binary outcomes. The effects of the different supplementation groups compared to daily placebo intake (reference) across follow-up visits were assessed by incorporating the intake groups and follow-up visits as indicator variables in the models. In addition, to assess the effects at each follow-up timepoint, we repeated the analyses with each follow-up visit serving as a reference group for all outcomes. To account for the correlation between the repeated measures within each participant, we included a random slope and grouping-factor for participants in the LMM and GEE analyses, respectively. Due to skewed data distributions, ferritin and gastrointestinal discomfort data were log transformed. Furthermore, each model was adjusted for the baseline proportions or levels of its respective dichotomous or continuous outcome variable. In addition to the crude models, adjusted analyses were performed in which sex, age, body mass index (BMI), self-reported infectious disease (including COVID-19), donation frequency (number of donations in the last two years), and treatment adherence were included in the models. Additionally, sensitivity analyses were performed using the 60 mg daily iron supplementation group as a reference, in order to differences between the iron supplementation groups. Effect modification by sex, age (<50 vs. ≥50 years), BMI (<25 vs ≥25 kg/m^2^), menstrual status (regularly menstruating, no menstruation due to contraception, no menstruation due to menopause, or no menstruation due to other reason, and donation frequency (<5 vs ≥5 donations in the two years prior to participating) was evaluated for the primary outcomes. A two-sided p-value of <0.05 is considered statistically significant. The statistical analyses were performed in R using RStudio (v4.0.3, RStudio, PBC, Boston) and the code used is available on GitHub (https://github.com/JKarregat/FORTE).

### Deviations from the protocol

Approximately 600 donors who expressed their willingness to participate in the trial did not visit a donation centre during the course of the study, as they were not routinely invited for a whole blood donation during that time. This led to inclusion of 830 participants in the trial rather than the desired 1200 based on the performed power calculation^12^. With only nine non-western European donors, ethnicity was not evaluated as a potential confounder due to limited ethnic diversity in our study population and consequent constraints on statistical power. Likewise, the prevalence of PICA (the consumption of non-food items) was insufficient for statistical evaluation within each supplementation group, with no more than four participants in total reporting PICA at any time point.

## RESULTS

Of the 15,780 invited donors, 2,676 (17%) expressed their willingness to participate and 2,052 subsequently attended a donation center where they provided their informed consent and donated whole blood. Ultimately, 830 donors with ferritin levels ≤30 µg/L were enrolled in the trial (Figure 1). Following a double-blinded, randomization procedure stratified by sex and age (≤50 and >50 year), between 135 to 140 participants were assigned to each of the trial arms.

**Figure 1.**
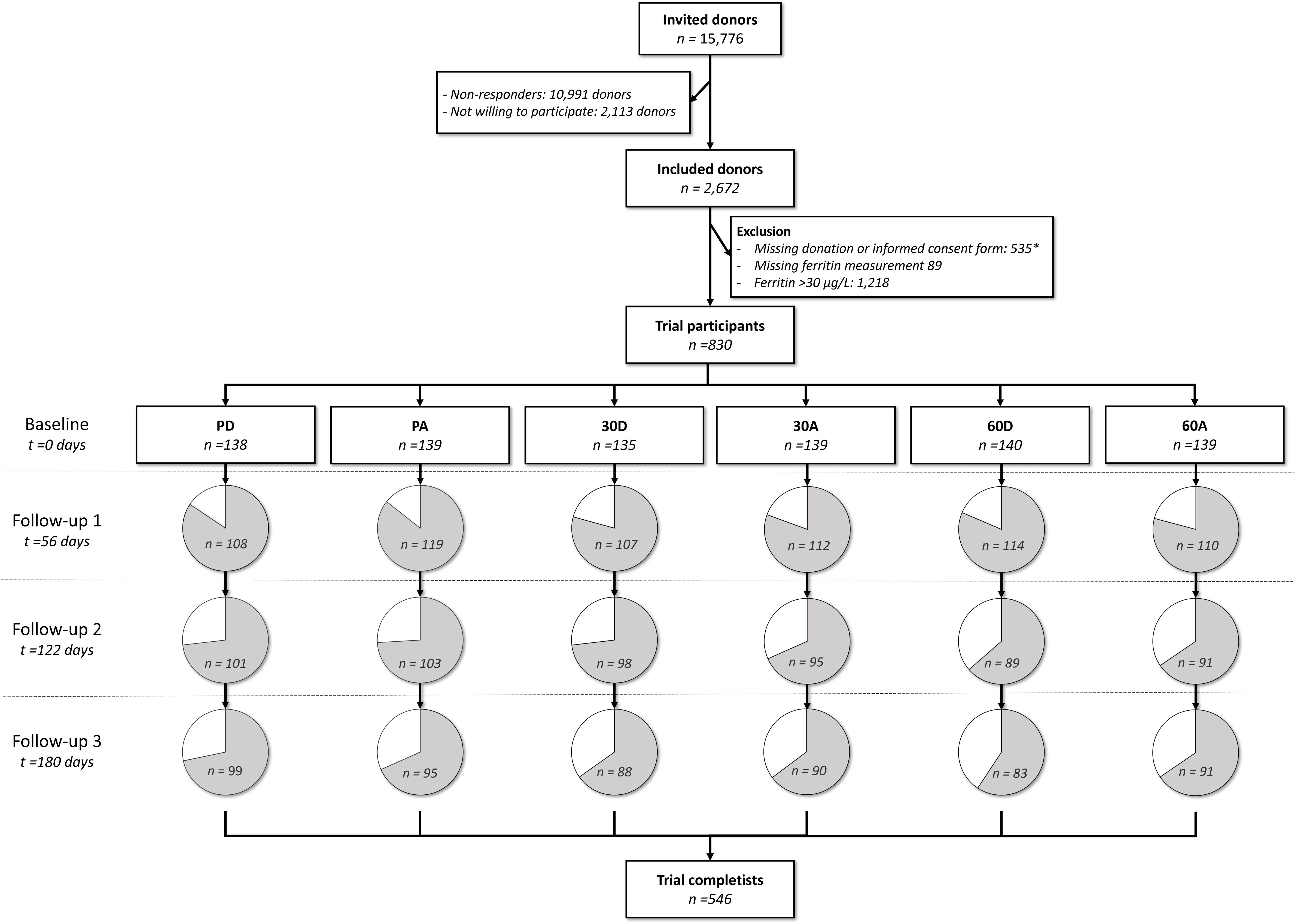
Schematic overview of the participant inclusion and follow-up process. PA: placebo alternate days, PD: placebo daily, 30A: 30mg alternate days, 30D: 30 mg daily, 60A: 60mg alternate days, 60D: 60mg daily. *Donors were not invited for a regular whole blood donation within the study period or did not provide their informed consent.

Baseline characteristics were well-balanced across the randomization groups (Table 1). Irrespective of group allocation, distribution of men and women, age, BMI, median ferritin levels, and mean Hb levels, number of donations in the past 2 years, prevalence of ID, and the proportion of used supplements were similar. However, the proportion of smokers was higher in the PA, 60D, and 60A-groups.

**Table 1.** Baseline participant characteristics stratified by supplementation group.

|  | PD | PA | 30D | 30A | 60D | 60A |
| --- | --- | --- | --- | --- | --- | --- |
| **n** | 138 | 139 | 135 | 139 | 140 | 139 |
| **Women (%)** | 80 (58.0) | 76 (54.7) | 77 (57.0) | 77 (55.4) | 78 (55.7) | 76 (54.7) |
| **Age (mean (SD))** | 42.4 (15.1) | 42.4 (15.8) | 42.2 (15.3) | 42.6 (15.1) | 41.7 (15.6) | 42.7 (15.5) |
| **BMI (mean (SD))** | 25.3 (5.0) | 24.8 (3.8) | 24.4 (3.00) | 24.8 (3.9) | 24.7 (3.6) | 24.6 (3.7) |
| **Number of donation in last 2 years (mean (SD))** | 5.0 (1.7) | 5.1 (1.6) | 5.1 (1.5) | 5.1 (1.5) | 5.1 (1.5) | 5.1 (1.8) |
| **Smoking (%)** | 3 (2.2) | 12 (8.6) | 5 (3.8) | 5 (3.7) | 8 (6.0) | 11 (8.0) |
| **Hb (mean (SD))** | 14.1 (1.0) | 14.0 (1.0) | 14.0 (1.6) | 14.0 (1.0) | 14.0 (1.0) | 14.0 (1.1) |
| **Ferritin (median [IQR])** | 21.0 [16.3, 25.0] | 20.0 [14.5, 26.0] | 20.0 [15.0, 25.0] | 21.0 [17.0, 25.0] | 21.0 [15.0, 27.0] | 20.0 [15.0, 25.0] |
| **Iron deficiency (%)** | 26 (18.8) | 35 (25.2) | 31 (23.0) | 28 (20.1) | 27 (19.3) | 29 (20.9) |
| **Low Hb (%)** | 7 (6.3) | 12 (11.0) | 9 (9.5) | 5 (4.4) | 12 (10.7) | 8 (7.6) |
| **Proportion of supplements used (mean (SD))\*** | 0.8 (0.1) | 0.8 (0.1) | 0.8 (0.2) | 0.8 (0.1) | 0.7 (0.2) | 0.8 (0.2) |

\*Data collected at 56 days.
PD: placebo daily, PA: placebo alternate days, 30D: 30 mg daily, 30A: 30mg alternate days, 60D: 60mg daily, 60A: 60mg alternate days

The proportion of participants with ID, low-ferritin, and low Hb at 56 days post-donation are presented in Figure 2, for each dosage and frequency of iron supplementation. Whereas the 48.7 and 50.9% of participants in the PD and PA-group presented with ID after the supplementation period, the 60D showed the lowest presence of ID with 0.9% of participants (Figure 2). Low Hb was present in more than 20% of both PD and PA-groups, whereas for the 60D group 1.8% of participants presented with low Hb (Figure 2). Compared to the PA-group, the proportion of participants with low ferritin was higher in both the 30A and 60A-groups, but compared to the PD-group, the 30D and 60D-groups had a lower presence of low ferritin.

**Figure 2.**
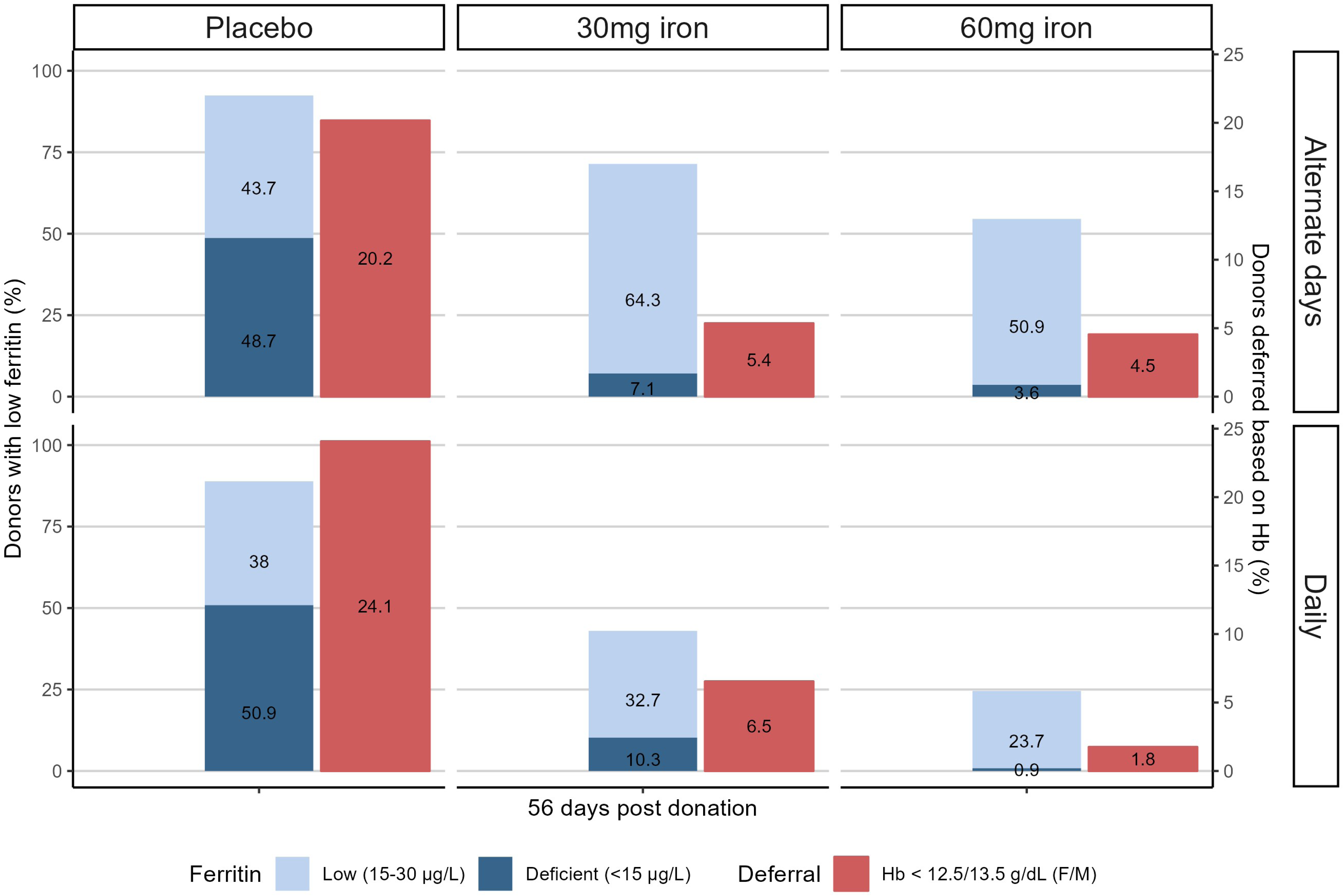
Proportion of participants with ferritin levels below the iron deficiency threshold (15 µg/L), low ferritin levels (ferritin 15-30 µg/L), and Hb levels below the Hb deferral cut-off (Hb <12.5 g/dL for women and Hb<13.5 g/dL for men) for each supplementation group at 56 days follow-up.

The effects of the different iron supplementation dosages and frequencies, compared to daily placebo supplementation, on ID, low-ferritin, and low Hb at 56 days post-donation are presented in Figure 3. All iron supplementation groups showed significantly lower odds of ID compared to the PD-group (reference group), with odds ratios (OR) ranging from 0.60 (95% CI: 0.55–0.66) for the 60D to 0.65 (95% CI: 0.59–0.72) for 60A-group (Figure 3). Similarly, for all iron supplementation groups, the odds of low ferritin after 56 days of supplementation were significantly lower in comparison to the reference group (Figure 3). For the 30D and 30A-groups, we observed ORs of 0.82 (95% CI [0.74-0.91]) and 0.70 (95% CI [0.63-0.78]), respectively, and for the 60D and 60A-groups, respectively, we observed ORs of 0.61 (95% CI [0.55-0.68]) and 0.52 (95% CI [0.47-0.57]) compared to the reference (Figure 3). Post-supplementation, the differences in primary outcomes between supplementation versus placebo groups became smaller over time, and but remained statistically significant (Appendix A1). We observed the same effect directions and patterns over time in ferritin as a continuous outcome (Appendix A1). Median ferritin trajectories per trial arm are presented in Appendix B1.

**Figure 3.**
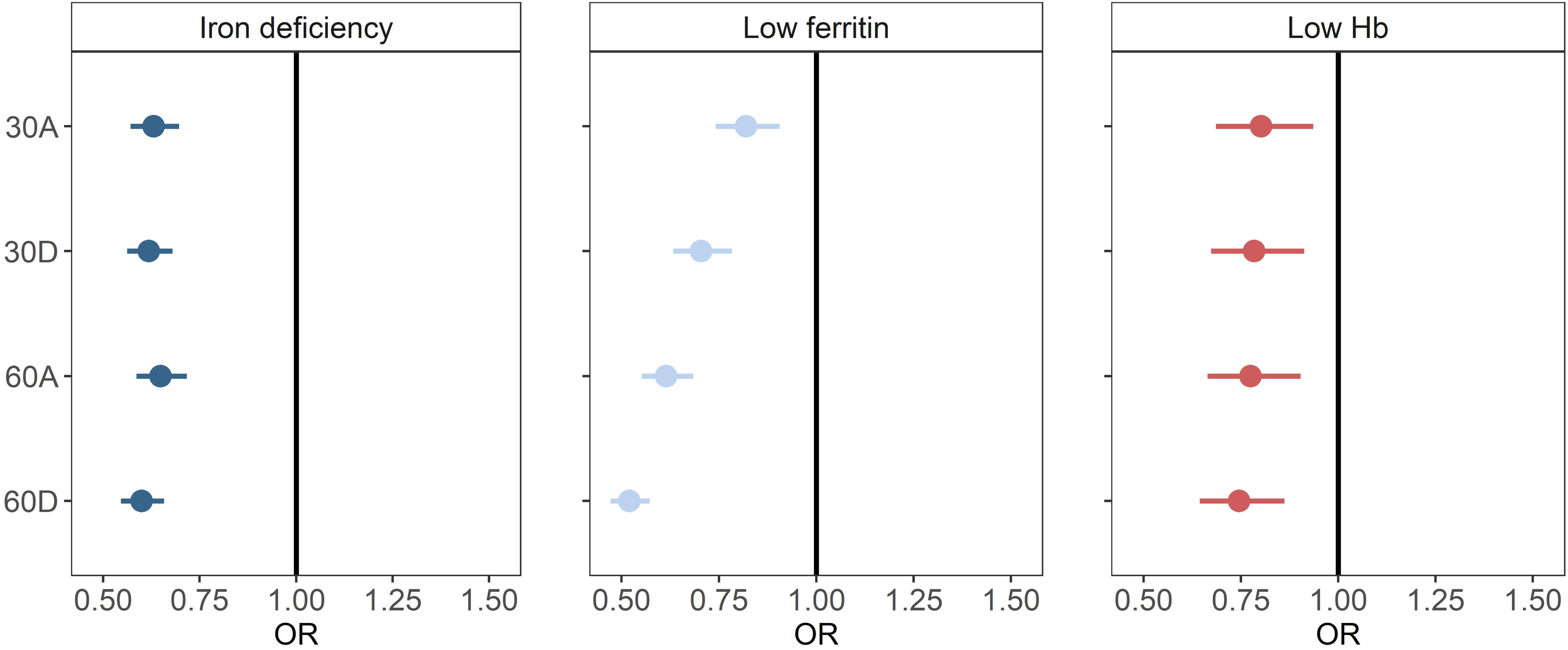
Generalized Estimating Equations (GEE) analyses showing the effects of iron supplementation compared to daily placebo supplementation at 56 days in terms of iron deficiency, low ferritin, and low Hb. All analyses were adjusted for baseline levels for each outcome. Filled circles represent a significant association. Iron deficiency is defined as ferritin levels <15 µg/L; low ferritin levels as ferritin 15-30 µg/L; and low Hb as Hb levels below the Hb deferral cut-off (Hb <12.5 g/dL for women and Hb<13.5 g/dL for men). 30A: 30mg alternate days, 30D: 30 mg daily, 60A: 60mg alternate days, 60D: 60mg daily

All iron supplementation frequencies and dosages were associated with lower odds of low Hb after 56 days, compared to placebo. The ORs were comparable, ranging from 0.74 (95% CI [0.64-0.86]) in the 30A to 0.80 (95% CI [0.68 - 0.93]) in the 60D-group. Post-supplementation, the ORs became smaller over time and were not significantly different from the OR in the reference group (Appendix A1). With Hb as a continuous outcome, we found approximately the same effect directions and patterns over time (Appendix A1). Mean Hb trajectories per trial arm are presented in Appendix B2.

For the primary outcomes, no substantial differences were observed for the adjusted models (Appendix A2) compared to the crude models. Effect modification was observed in terms of sex (Appendix C1) and the number of donations (Appendix C2) for ID, showing a stronger effect of iron supplementation on ID for women and donors with fewer than five donations in the two years prior to participating compared to men and donors with 5 or more donations, respectively. A stronger effect of iron supplementation on low Hb was observed for donors age 18-49 years compared to donors 50 years and older (Appendix C3).

Figure 4 shows the effects of iron supplementation dosage and frequency in terms of general health, fatigue, cognitive failure, restless legs syndrome, gastrointestinal discomfort, intention to return to donate, and treatment adherence, compared to the PD-group. No significant differences with the reference group were observed in terms of general health, fatigue, cognitive failure, and RLS (Figure 4). Likewise, no consistent significant patterns were observed for the sub scores for mental and physical health (Appendix B1). Iron supplementation was not associated with self-reported gastrointestinal discomfort overall. However, PA and 60D led to a factor 1.16 (95% CI [1.01-1.34]) and 1.22 (95% CI [1.06-1.41]) higher score, respectively, for self-reported constipation compared to the PD-group at 56 days, respectively (Appendix C). Intention to return to donate was not significantly different for the iron supplementation groups compared to PD-group at any time-point. However, in all groups except for the 60D-group, intention to return to donate was lower at the follow-up visits compared to baseline (Appendix F6). The only significant difference in treatment adherence compared to the PD-group was observed in the 60A-group, where adherence was 6% higher (0.06, 95% CI [0.01-0.11]).

**Figure 4.**
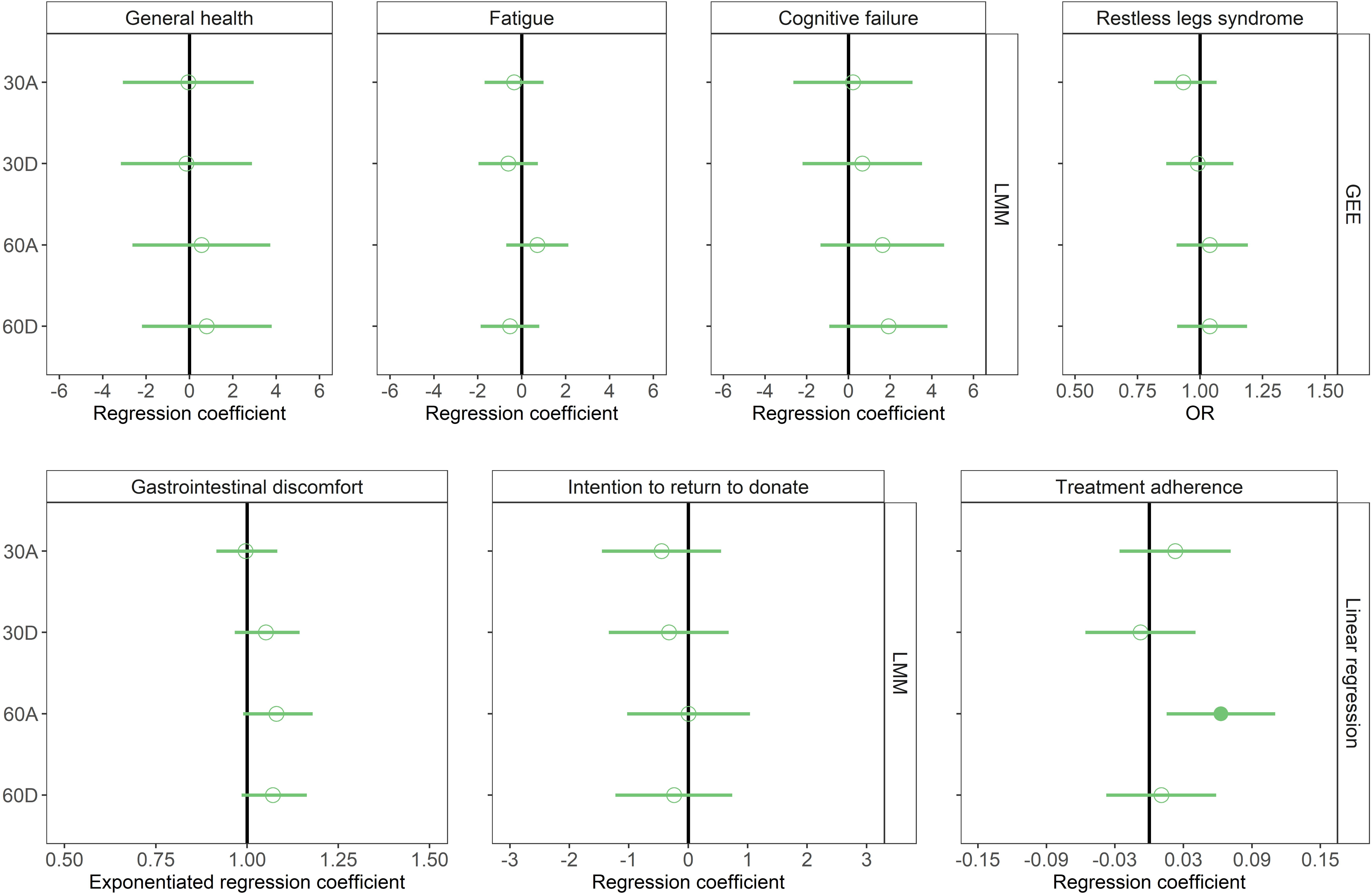
Generalized Estimating Equations (GEE), Linear Mixed Models (LMM), and linear regression analyses showing the effects of iron supplementation and alternate day placebo supplementation compared to daily placebo supplementation at 56 days follow-up in terms of general health, fatigue, cognitive failure, restless legs syndrome, gastrointestinal discomfort, intention to return to donate, and treatment adherence. All analyses were adjusted for baseline levels for each outcome. Filled circles represent a significant association. 30A: 30mg alternate days, 30D: 30 mg daily, 60A: 60mg alternate days, 60D: 60mg daily

## DISCUSSION

This randomized trial demonstrates that all examined dosages and frequencies of iron supplementation effectively reduce iron deficiency, low ferritin levels, and low hemoglobin levels in regular whole blood donors with low ferritin levels. Specifically, daily supplementation with 60 mg of elemental iron emerges as the optimal strategy, as it significantly reduces the odds of low ferritin levels without any notable differences in the odds of iron deficiency, low hemoglobin, and side effects, or treatment adherence compared to other supplementation strategies.

Our findings of a reduced risk of ID, low ferritin, and low Hb following iron supplementation in regular donors align with those of previous studies^7,9,19^. However, this is the first study to evaluate both iron dosage and intake frequency in relation to iron stores and donor health, while also considering side effects, intention to return to donate, and treatment adherence. Difference in dose- and intake frequency response for low ferritin levels might be related to the acute hepcidin release following iron supplementation, which inhibits iron absorption in the intestine.^8,10,20^ Galetti *et al.* recently demonstrated a linear relationship between ferritin and hepcidin, indicating a decrease in iron uptake from 21.9% to 10.0% as ferritin levels increased from 1 µg/L to 30 µg/L.^21^ This decrease in the proportion of iron taken up with increasing ferritin levels, could explain why higher dosed iron supplements taken more frequently are more effective at addressing low ferritin levels (ferritin ≤ 30 µg/L) compared to low-dosed supplements taken less frequently.

Although several clinical studies have shown an association between iron supplementation-related gastrointestinal side effects and reduced treatment adherence, we observed high adherence, likely due to minimal reporting of gastrointestinal discomfort.^22–24^ Previous research by Tolkien *et al.,* demonstrated that iron supplementation in the form of ferrous sulphate led to over twice the odds of experiencing gastrointestinal side effects compared to placebo.^25^ The lower levels of gastrointestinal discomfort observed in the present study may be attributed to the use of ferrous bisglycinate, an iron formulation that includes iron-bound chelates, which prevent binding to absorption-inhibiting dietary compounds and thereby result in higher fractional iron uptake.^26–28^ This, as well as the higher level of intestinal absorption compared to other iron salts, lowers the amount of iron entering the colon and in turn lowers gastrointestinal side-effects.^29,30^ These findings further highlights the potential of ferrous bisglycinate as a low dosed iron formulation for ferritin-guided iron supplementation in donors.

No improvements in ID-related symptoms were observed across any of the iron supplementation groups compared to placebo. Studies evaluating improvements after iron supplementation in ID-related symptoms in non-anemic iron deficient individuals provide conflicting results.^19,31–34^ Previous research in Dutch donors suggested that the positive feelings associated with donating can offset potential negative effects, which might have led to an overall underreporting of ID-related symptoms.^35^ No differences in the intention to return to donate were observed across any of the iron supplementation groups compared to the placebo group at any timepoint. These findings are in line with results from a cross-sectional survey study, evaluating the perceptions of Dutch whole blood donors on post-donation iron supplementation.^36^ This study showed that only 7% of the questioned donors indicated that iron supplementation as a blood service policy would have a negative effect on their motivation to donate. However, the absence of any association between iron supplementation and the intention to return to donate might be attributable to self-selection bias. Donors willing to participate in this randomized trial study were likely to have had some degree of openness towards iron supplements. The effect of iron supplementation as a policy on donor return should therefore be further evaluated through an implementation study.

Strengths of this study include the large study sample and comprehensive evaluation of both biochemical and health outcomes including symptom relieve, potential side effects of iron supplementation, and intentions to donate. This allowed us to evaluate how iron supplementation, if implemented in a blood service, might impact both donor health and intention, and consequently donor availability. Data on ID-related symptoms were retrieved using self-reported surveys, and might therefore lack sensitivity to evaluate subtle changes in these non-specific symptoms (i.e., not specifically related to one medical condition) in a relatively healthy donor population^37^. Although about one-third of the participants were lost to follow-up, the distribution of attrition was similar across all supplementation groups. Furthermore, robust analysis methods (i.e., GEE and LMM), were used to effectively handle missing data, allowing us to include all available data points regardless of missing data at previous or subsequent study visits.

Iron supplementation results in shorter recovery periods and would allow donors to continue to donate regularly compared to deferral. Previous research has shown that ferritin-guided deferral decreases donor return and, in turn, donor availability.^5^ In addition, one-third of the donors indicated in a survey study to equally prefer iron supplementation or deferral as an iron management policy.^36^ Iron supplementation could therefore be implemented as a policy on its own or alongside ferritin-guided deferral, to improve donor availability. Following the Dutch guidelines for ferritin-based deferral, we used ferritin levels below 30 µg/L as a criterium to include donors in the study. However, due to the limited harmonization of ferritin measurements internationally, this cut-off is not transferable and thus blood services should define their own cut-off for supplementation^38,39^.

Based on the current findings, 60 mg of daily iron supplementation will be considered by the medical advisory board of Sanquin for implementation as a blood service policy. This dosage and frequency was shown to be effective for improving iron deficiency, low ferritin, and low Hb, was well-tolerated in terms of gastrointestinal side-effects, and did not affect intention to return to donate. Our findings could guide the decision making of blood services on iron supplementation policies to protect their donors from developing iron deficiency and enhance donor availability.

## Data Availability

All data produced in the present study are available upon reasonable request to the authors.

