## Supplement for "Ferritin-guided iron supplementation in whole blood donors (FORTE): results of a double-blind randomized controlled trial"

### Appendix A

Adjusted Generalized Estimating Equations (GEE) and Linear Mixed Models (LMM) analyses for the primary outcomes did not result in substantial differences compared to the unadjusted models. However, for Hb at 180 days the regression coefficient of the 60D-group was no longer significant.

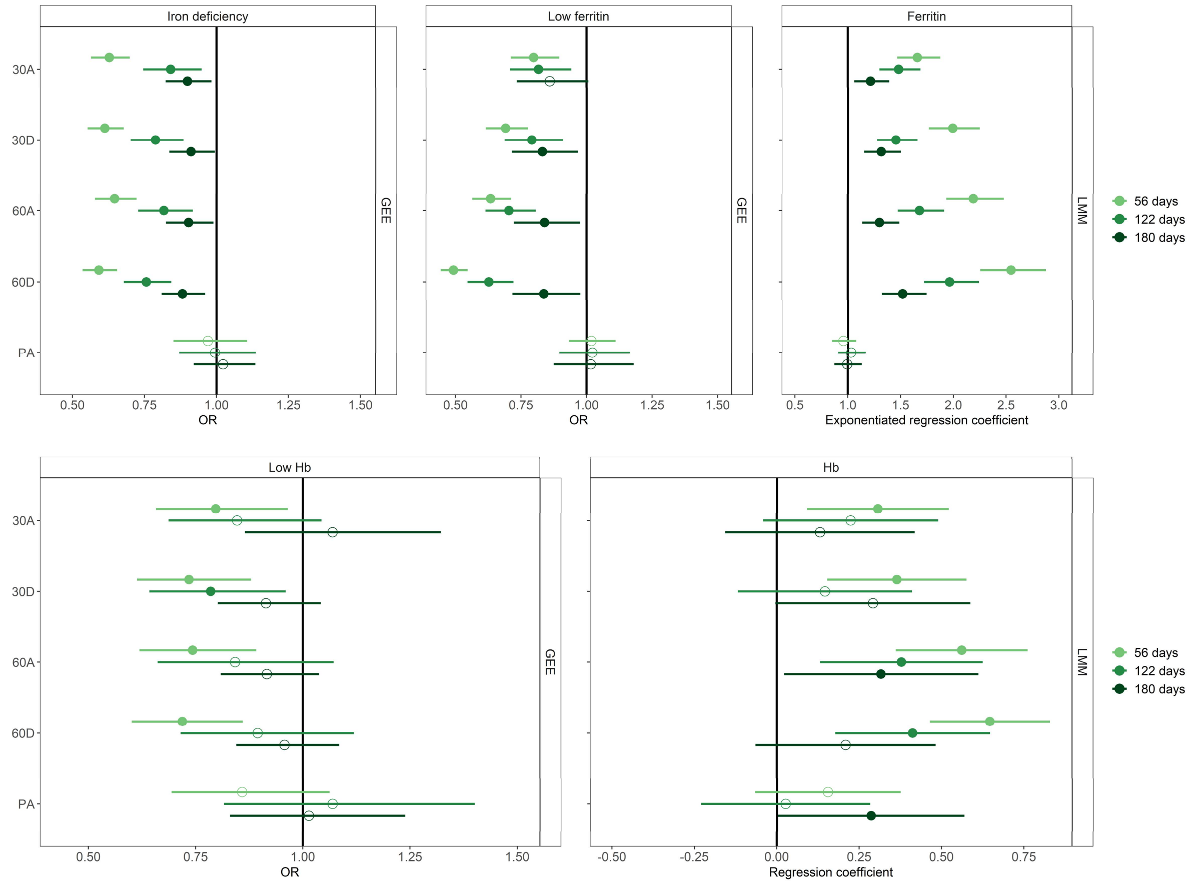

***Appendix A1.*** *Adjusted Generalized Estimating Equations (GEE) and Linear Mixed Models (LMM) analyses showing the effects of iron supplementation and alternate day placebo supplementation compared to daily placebo supplementation for the primary outcomes at each timepoint. All models were adjusted for sex, age, BMI, recent infectious disease, number of donation in the two years prior to baseline donation, and the proportion of used supplements. Filled circles represent a significant association. 30A: 30mg alternate days, 30D: 30 mg daily, 60A: 60mg alternate days, 60D: 60mg daily, PA: placebo alternate days.*

Generalized Estimating Equations (GEE) and Linear Mixed Models (LMM) analyses showed that the estimated effects converged towards the reference group over time. At 180 days, the OR for iron deficiency and low ferritin and the exponentiated regression coefficient for ferritin remained significantly distinct for all iron supplementation groups compared to placebo. For low Hb and Hb, only the 60D-group group showed significantly higher regression coefficients at 180 days for Hb compared to the PD-group.

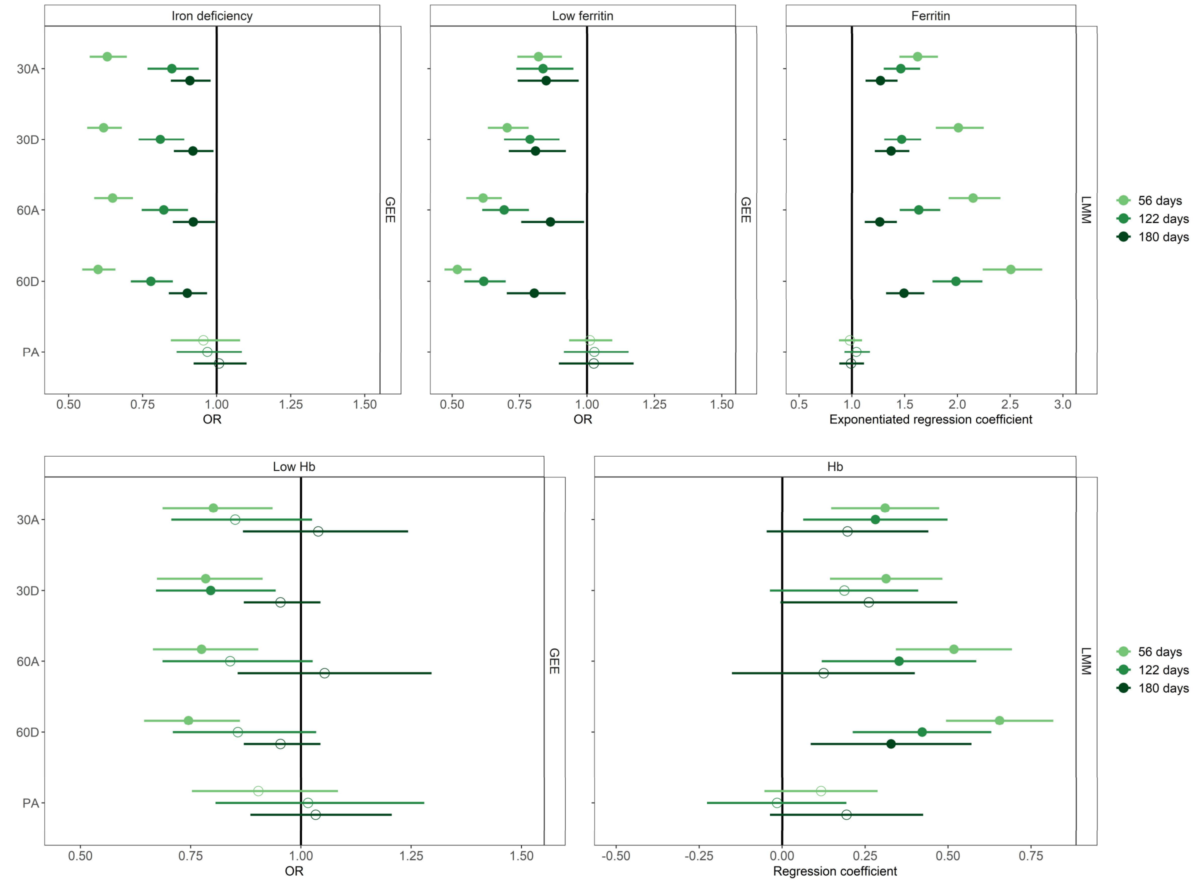

***Appendix A2.*** *Generalized Estimating Equations (GEE) and Linear Mixed Models (LMM) analyses showing the effects of iron supplementation and alternate day placebo supplementation compared to daily placebo supplementation for the primary outcomes at each timepoint. Filled circles represent a significant association. 30A: 30mg alternate days, 30D: 30 mg daily, 60A: 60mg alternate days, 60D: 60mg daily, PA: placebo alternate days.*

### Appendix B

Median ferritin level trajectories (B1) were similar for the placebo supplementation groups, exhibiting a decrease at 56 days compared to baseline levels, followed by a steady increase until 180 days. In contrast, median ferritin levels were higher at 56 days compared to baseline levels in all iron supplementation groups, with steeper increases in the daily intake groups compared to alternate days intake. Post-supplementation median ferritin level trajectories were slightly different across the iron supplementation groups, with a steady increase in the alternate days intake groups and a flattening increase for the daily intake groups.

Mean Hb levels (B2) exhibited similar patterns for the placebo supplementation groups for both men and women, showing a decrease at 56 days compared to baseline levels, followed by a steady increase until 180 days. Mean Hb trajectories were similar for men and women in terms of change over time within each iron supplementation group. However, whereas mean Hb levels increased after 56 days of daily iron supplementation for both 30- and 60mg intake, mean Hb levels remained stable in the alternate days iron supplementation groups. A steady increase in mean Hb levels was observed in all iron supplementation groups during the post-supplementation period.

**1**

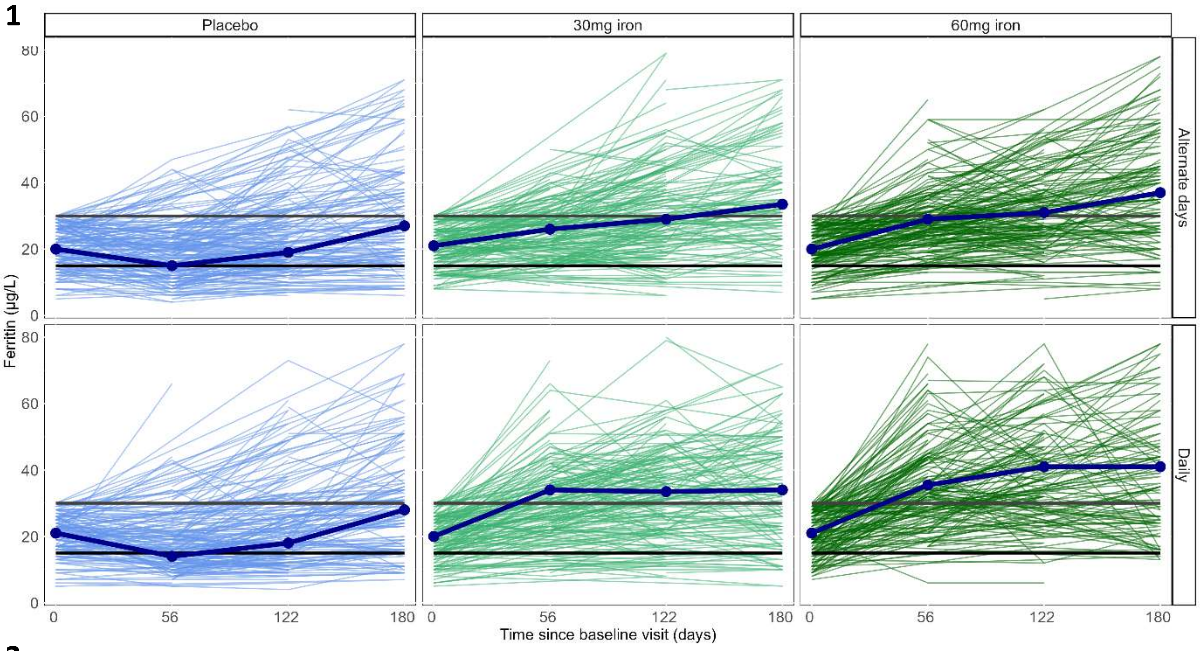

**2**

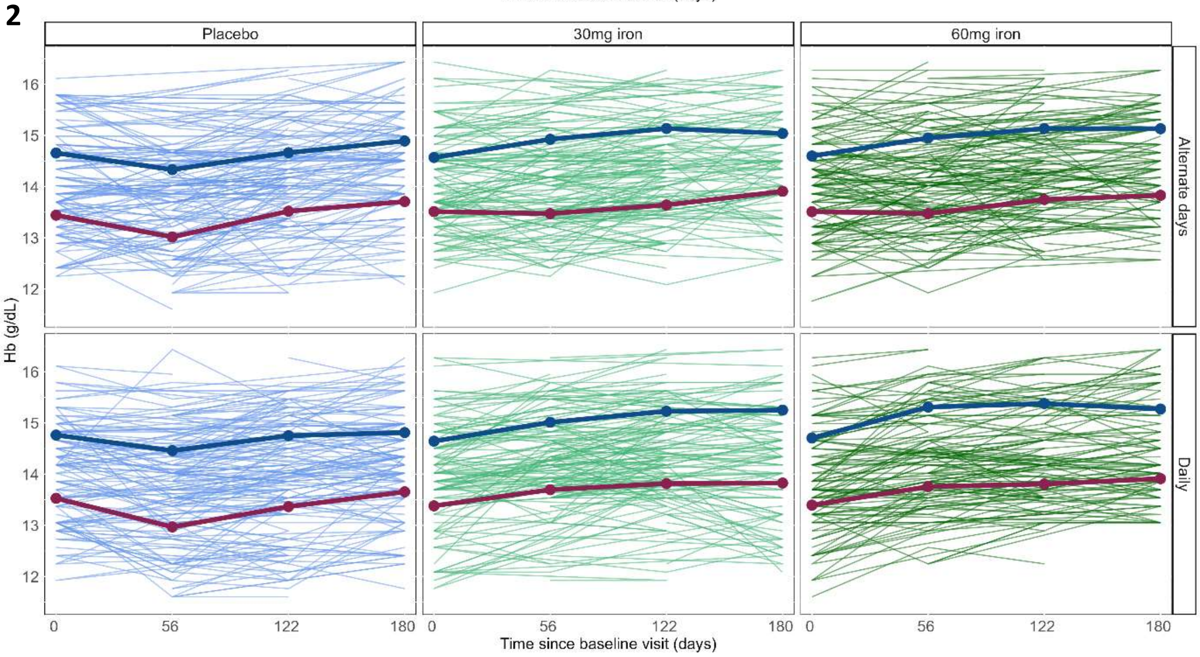

***Appendix B.*** *Trajectories of ferritin- (1) and Hb levels (2), for each supplementation separately. Median ferritin trajectories are indicated by the dark blue line. Mean Hb level trajectories for men are indicated by the dark blue line and for women by the dark red line.*

#### Appendix C

At 56 days, odds of iron deficiency for men (Appendix C1) were similar and ranged from 0.72 (95% CI [0.63-0.82]) for the 30A-group to 0.74 (95% CI [0.65-0.84]) for the 30D-group, compared to PD-group (reference). For women, odds of iron deficiency at 56 days ranged from 0.52 (95% CI [0.46-0.59]) for 60D-group to 0.59 (95% CI [0.51-0.67]) for the 60A-group, compared to the reference group. Whereas, the odds of iron deficiency remained significantly lower for all iron supplementation at 122 days for women compared to the reference group, for men only the 60A- and 60D-groups showed significantly lower odds. At 180 days, for both men and women, odds of iron deficiency were not significantly different for the iron supplementation groups compared to the reference group.

At 56 days, odds of iron deficiency for donor with 0-4 donations in the two years prior to the baseline donation (Appendix C2) ranged from 0.48 (95% CI [0.41-0.56]) for the 60D-group to 0.60 (95% CI [0.50-0.71]) for the 60A-group, compared to the reference group. For donor with 5 or more donation, the odds of iron deficiency at 56 days ranged from 0.52 (95% CI [0.46-0.59]) for 60D-group to 0.59 (95% CI [0.51-0.67]) for the 60A-group, compared to the reference group. The odds for iron deficiency were not significantly different across all iron supplementation groups compared to the reference group at 180 days for donors with more than prior 5 donations. In contrast, the odds remained significantly lower for the iron supplementation groups for donors with 0-4 prior donations, apart from 30A-group.

At 56 days, odds of low Hb were for donor between 18 and 49 years of age were lower for the iron supplementation groups, apart from 30A-group, compared to the reference group (Appendix C3). Odds ranged from 0.72 (95% CI [0.60-0.86]) for the 60A-group to 0.76 (95% CI [0.63-0.91]) for the 30D-group. Odds returned to non-significant levels at 122 and 180 days. No significant differences between the iron supplementation groups and the reference groups in terms of low Hb was observed for donors of 50 years and older. At 180 days, the odds for low Hb in the donors aged 18-49 years group could not be assessed for the 60A- and 30D- and 60D-groups, as no donors low Hb were present.

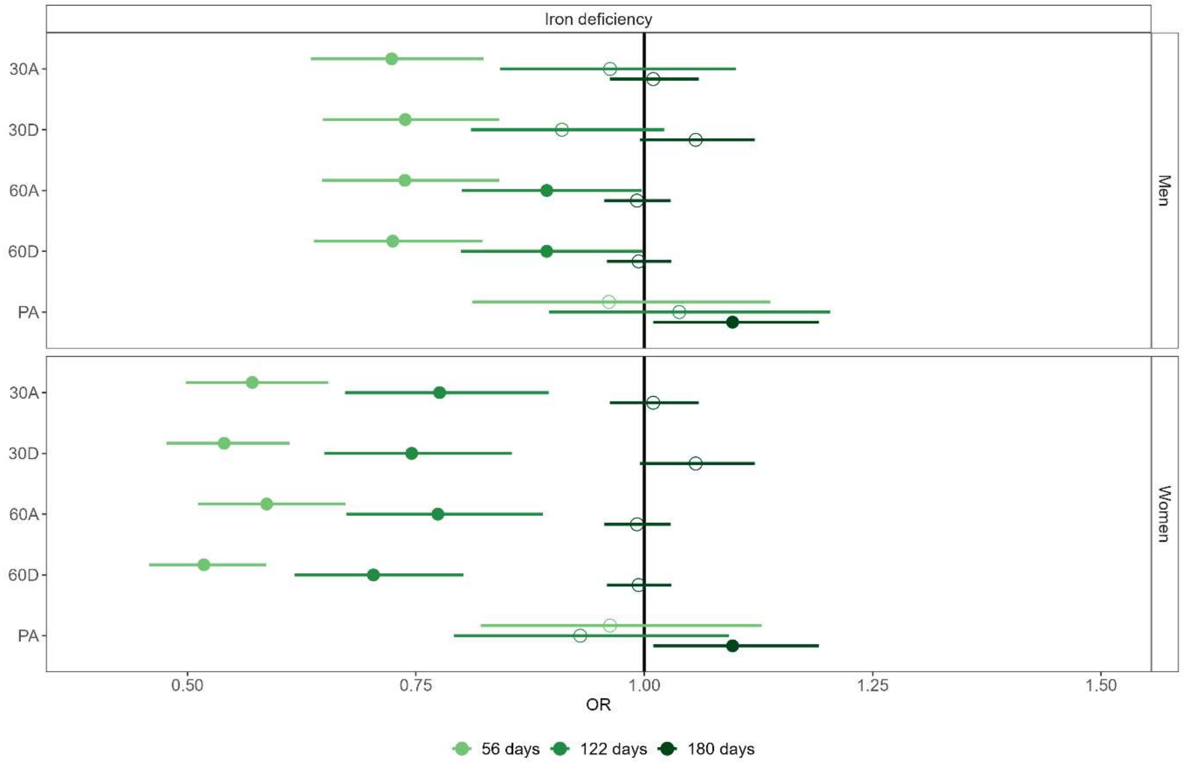

***Appendix C1***. *Generalized Estimating Equations (GEE) analyses showing the effects of iron supplementation and alternate day placebo supplementation compared to daily placebo supplementation for iron deficiency at each timepoint, for men and women seperately. Filled circles represent a significant association. 30A: 30mg alternate days, 30D: 30 mg daily, 60A: 60mg alternate days, 60D: 60mg daily, PA: placebo alternate days.*

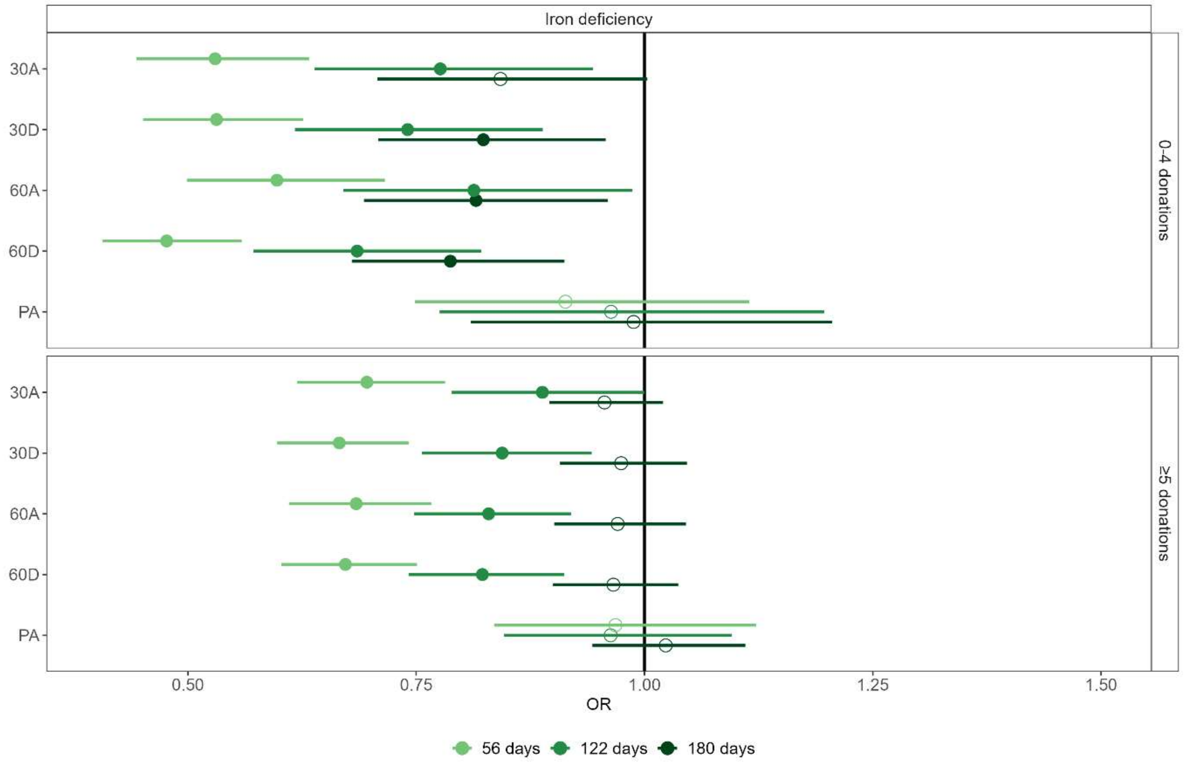

***Appendix C2***. *Generalized Estimating Equations (GEE) analyses showing the effects of iron supplementation and alternate day placebo supplementation compared to daily placebo supplementation in terms of iron deficiency at each timepoint, for donors with 0-4 donations and ≥5 donation in the two years prior to the baseline donation seperately. Filled circles represent a significant association. 30A: 30mg alternate days, 30D: 30 mg daily, 60A: 60mg alternate days, 60D: 60mg daily, PA: placebo.*

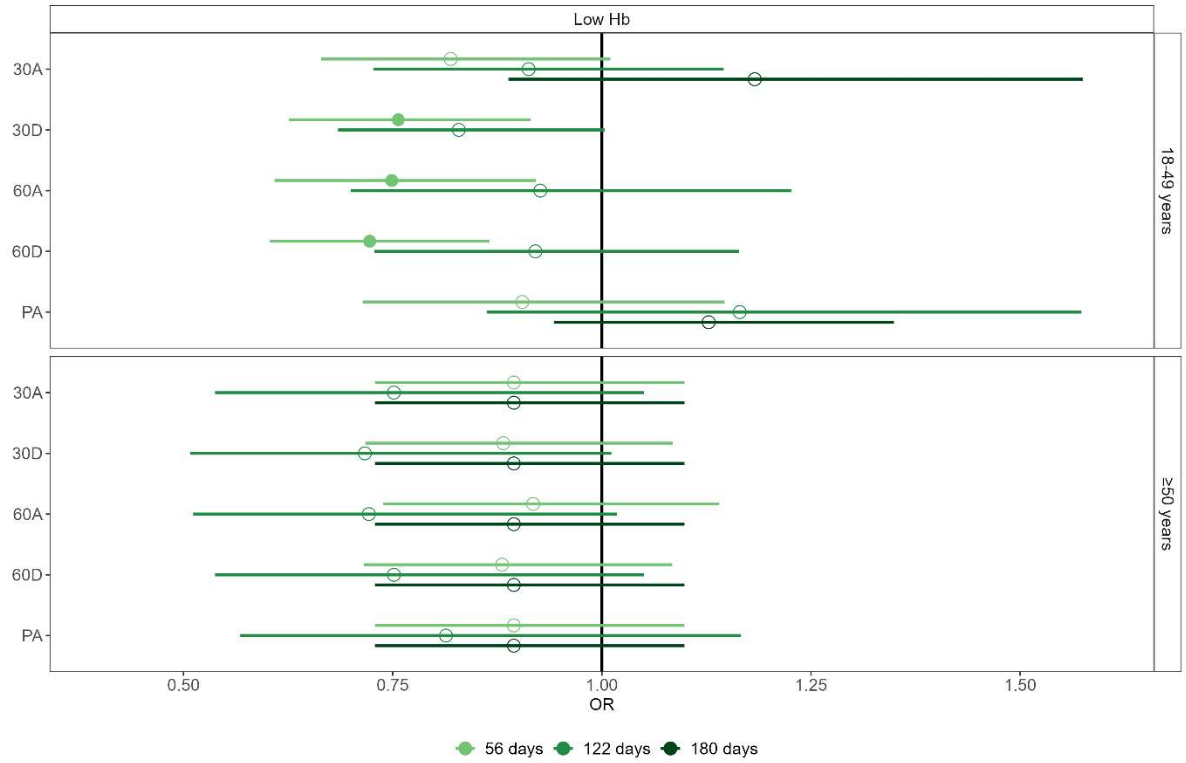

***Appendix C3.*** *Generalized Estimating Equations (GEE) analyses showing the effects of iron supplementation and alternate day placebo supplementation compared to daily placebo supplementation in terms low Hb levels at each timepoint, for donors 18-49 years and ≥50 years of age seperately. Filled circles represent a significant association. 30A: 30mg alternate days, 30D: 30 mg daily, 60A: 60mg alternate days, 60D: 60mg daily, PA: placebo alternate days.*

### Appendix D

No differences in terms of physical and mental health, physical fatigue, and the gastrointestinal sub scores diarrhoea, abdominal pain, reflux, and indigestion, was observed between all iron supplementation- and alternate day placebo groups compared to daily placebo supplementation groups. At 180 days, 30A-group was associated with a lower mental fatigue score (β = 1.29 (95% CI [-2.40—0.18]) compared to PD-group. At 56 days, the PA- and 60D-groups was showed an 1.16 (95% CI [1.01-1.34] and 1.22 (95% CI [1.06-1.41] times higher scores for constipation compared to daily placebo supplementation, respectively.

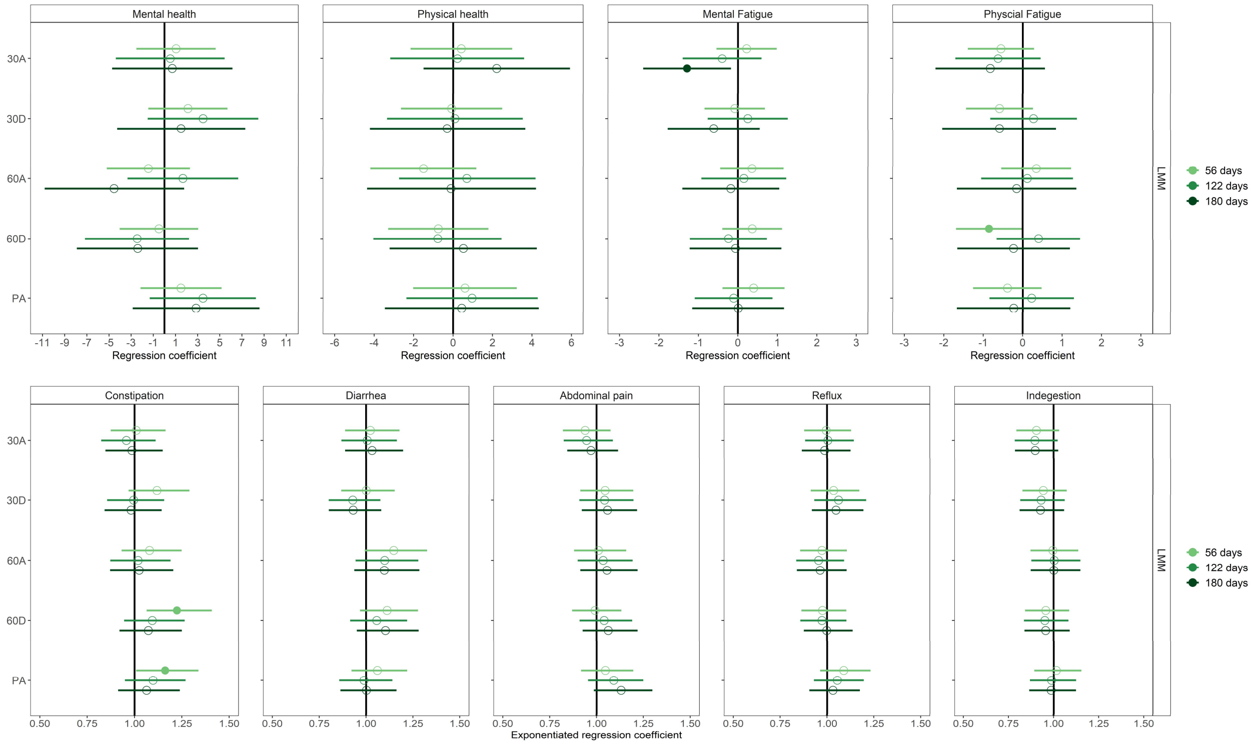

***Appendix D.*** *Linear Mixed Models (LMM) analyses showing the effects of iron supplementation and alternate day placebo supplementation compared to daily placebo supplementation for the subscores of the secondary outcomes at each timepoint. Filled circles represent a significant association. 30A: 30mg alternate days, 30D: 30 mg daily, 60A: 60mg alternate days, 60D: 60mg daily, PA: placebo alternate days.*
